## Supplementary Materials 1 for "Complex interventions for aggressive challenging behaviour in adults with intellectual disability: a rapid realist review informed by multiple populations"

**Supplementary Table 1** Search strategy for MEDLINE, EMBASE, PsycINFO and HMIC using the Ovid interface on 27/07/2020

| Population of interest | Alzheimer* OR Angelman OR angelman syndrome OR ASD OR Asperger* syndrome OR autis* disorder* OR autis* spectrum condition* OR autis* spectrum disorder OR complex need* OR complex support need* OR cornelia de lange OR cri du chat OR DD OR de lange OR Dementia OR development* delay* OR development* difficult* OR developmental disabilit* OR developmental disorder* OR developmental* impair* OR down* syndrome OR externali?ing disorder* OR externali?ing problem* OR fragile x OR fragile x syndrome OR genetic disorder* OR high support need* OR ID OR IDD OR intellectual development* disab* OR intellectual development* disorder* OR intellectual* deficien* OR intellectual* disab* OR intellectual* impair* OR klinefelter syndrome OR LD OR intellectual deficien* OR intellectual difficult* OR intellectual disab* OR intellectual impair* OR mental* deficien* OR mental* handicap* OR mental* health OR mental* illness* OR mental* retard* OR neurodevelopmental disabilit* OR neurodevelopmental disorder* OR PDD OR PDD-NOS OR pervasive developmental* disorder* OR pervasive developmental* disorder* otherwise specified OR PIMD OR PMLD OR prader willi syndrome OR profound* intellectual* multiple disab* OR profound* multiple intellectual disab* OR Rett syndrome OR SEN OR SEND OR smith magenis OR special education* need* disabilit* OR special education* need* OR special need* disabilit* OR special need* OR velocardiofacial syndrome OR william syndrome |
| --- | --- |
| Topic of interest | Aggression OR aggressive behavio?r* OR agitation OR anger OR behavio?ral problem* OR challenging behavio?r* OR disruptive behavio?r* OR disruptive disorder OR externali?ing behavio?r* OR frustration OR hostile behavio?r* OR hostility OR IED OR intermittent explosive disorder OR irritability OR mood dysregulation OR problem* behavio?r* OR rage OR violence OR violent behavio?r* |
| Programme of interest | ABA OR active support OR anger control training OR anger management OR applied behavio?r analysis OR care package* OR CBT OR cognitive behavio?r* therap* OR cognitive behavio?r* treatment* OR cognitive control training OR complex approach* OR complex intervention* OR complex program* OR complex strateg* OR complex therap* OR complex training* OR complex treatment* OR DBT OR dialectical behavio?r* therap* OR dialectical behavio?r* treatment* OR manuali?ed approach* OR manuali?ed intervention* OR manuali?ed program* OR manuali?ed strateg* OR manuali?ed therap* OR manuali?ed training* OR manuali?ed treatment* OR mindfulness OR multidisciplinary approach* OR multidisciplinary intervention* OR multidisciplinary program* OR multidisciplinary strateg* OR multidisciplinary therap* OR multidisciplinary training* OR multidisciplinary treatment* OR multimodal approach* OR multimodal intervention* OR multimodal program* OR multimodal strateg* OR multimodal therap* OR multimodal training* OR multimodal treatment* OR nonpharmacolog* approach* OR nonpharmacolog* intervention* OR nonpharmacolog* program* OR nonpharmacolog* strateg* OR nonpharmacolog* therap* OR nonpharmacolog* training* OR nonpharmacolog* treatment* OR PBS OR personali?ed approach* OR personali?ed intervention* OR personali?ed program* OR personali?ed strateg* OR personali?ed therap* OR personali?ed training* OR personali?ed treatment* OR positive behavio?r* support OR psychological approach* OR psychological intervention* OR psychological program* OR psychological strateg* OR psychological therap* OR psychological training* OR psychological treatment* OR psychosocial approach* OR psychosocial intervention* OR psychosocial program* OR psychosocial strateg* OR psychosocial therap* OR psychosocial training* OR psychosocial treatment* OR self-control training OR sensory approach* OR sensory intervention* OR sensory program* OR sensory strateg* OR sensory therap* OR sensory training* OR sensory treatment* OR social approach* OR social intervention* OR social program* OR social strateg* OR social therap* OR social training* OR social treatment* |
