## Supplementary Materials 2 for "Complex interventions for aggressive challenging behaviour in adults with intellectual disability: a rapid realist review informed by multiple populations"

| **Supplementary Table 2**  Summary of studies included in the rapid realist review | | | | | | |
| --- | --- | --- | --- | --- | --- | --- |
| **No** | **Author (date)** | **n** | **Population/ sample** | **Study design** | **Country** | **Name and description of intervention** |
| 1 | Ahemaitijiang et al. (2019) | 3 | Mothers of Chinese adolescents with autism spectrum disorder and aggression | Single-subject experimental design: Multiple baseline | China | Soles of the Feet (SoF) Mindfulness - Mothers were taught a basic foundational meditation practice (two 30-min sessions), followed by instructions in the SoF practice (four 30 min sessions). Once proficient in these two practices, the mothers taught their children to use SoF to reduce aggressive behaviours. |
| 2 | Appelhof et al. (2019) | 274 | Adults with young onset dementia (below the age of 65) | Randomised controlled trial, stepped wedge cluster design | The Netherlands | Grip on Neuropsychiatric symptoms in Institutionalized People with Young-onset Dementia - The group intervention consisted of an educational program combined with a care program for multidisciplinary teams involved with caring for people. Included evaluation of psychotropic drug prescription, detection, analysis, treatment, and evaluation of treatment of neuropsychiatric symptoms. Follow-up assessments were conducted for each group every 6 months for 18 months. |
| 3 | Ballard et al. (2020) | 847 | Older adults with Dementia | Cluster randomised controlled trial | UK | Improving Wellbeing and Health for People with Dementia (WHELD) – a person-centred care and psychosocial intervention, training staff in care homes to deliver manualised sessions. This included writing strength based care plans, providing structured social activities and setting individual goals. 60 minutes per week per person was allocated for a period of 9 months. |
| 4 | Ballard et al. (2009) | 318 | Older adults with Alzheimer disease with clinical agitation (including aggression) | Feasibility study for a randomised blinded placebo-controlled trial | UK | Brief Psychosocial Therapy – 4 week psychological intervention delivered by trained therapists to paid carers. This individual therapy included a focus on social interaction, personalised music and removal of environmental triggers. |
| **No** | **Author (date)** | **n** | **Population/ sample** | **Study design** | **Country** | **Name and description of intervention** |
| 5 | Bambara et al. (2001) | 19 | Members of community based teams involved in the care of people with intellectual disabilities and history of challenging behaviour | Qualitative interviews and multisite design | USA | Positive Behavioural Support – the study explored how teams implement and understand the process of positive behaviour support in four residential community-based teams. |
| 6 | Benson et al. (1986) | 54 | Adults with mild to moderate intellectual disability and aggression | Non-randomised assignment to two intervention groups | USA | Cognitive-Behavioural Anger Management Intervention – one group practiced relaxation during roleplays of anger-arousing situations. A self-instruction group were taught problem-solving. Treatment groups met in 12 weekly 90 min sessions. |
| 7 | Borowsky et al. (2004) | 224 | Children with behavioural disorders (aged 7-15 years) | Randomised controlled trial | USA | Primary Care Based Intervention for Children’s Violent Behaviours and Violence-Related Injuries – included psychosocial screening and a telephone-based parenting education programme, Positive Parenting, which emphasises nurturance, discipline and granting of psychologic autonomy. This manualised programme consisted of 15-30 minute weekly sessions for 13 lessons. |
| 8 | Bowers et al. (2015) | 564 | Adults in acute psychiatric wards with a range of conditions | Pragmatic cluster randomised controlled trial | UK | Safewards – staff across 31 wards implemented a package of 10 Safewards interventions (e.g. de-escalation, soft-words, sensory modulation tools) for 8 weeks. |
| 9 | Bradshaw et al. (2004) | 60 | Staff (n=38) and adults with profound/severe intellectual disability (n=22) | Experimental design (3 community houses) | UK | Active Support –Training included classroom-based training and interactive training. The first 2 houses received group training over 2 consecutive days. The third house received 1 day training and 2 hour individual sessions for each staff member. Four additional sessions were also then held for all. |
| **No** | **Author (date)** | **n** | **Population/ sample** | **Study design** | **Country** | **Name and description of intervention** |
| 10 | Chilvers et al. (2011) | 15 | Female residents of a medium secure unit with intellectual disabilities | Repeated measures design | UK | Mindfulness-Based Intervention – received twice a week in a ward environment for 6 months. Sessions lasted for 30 minutes. |
| 11 | Cullen et al. (2012) | 84 | Male adult inpatients with psychiatric disorders and a history of violence | Randomised controlled trial | UK | Cognitive Skills Intervention - Reasoning and Rehabilitation (R&R) program. Manualised programme targeting social problem-solving skills and thinking styles. Delivered over a minimum of 36 2-hour sessions and includes 8 modules. |
| 12 | Davies et al. (2020) | 8 | Staff of adults with intellectual disability in a 7 bed assessment and treatment unit | Mixed methods design - repeated measures and qualitative study | UK | Safewards – staff were all trained in the 10 interventions and these were implemented 1 month at a time over 12 months. |
| 13 | Edwards et al. (2019) | 11 | Caregivers of children (aged 2-7 years) with autism spectrum disorder | Quantitative experimental (feasibility study) | USA | RUBI Parent Training for Disruptive Behaviours – manualised group intervention delivered to caregivers, consisting of 60-90 minute 11 core sessions, 1 individual telephone call and 2 assessment sessions (14 sessions in total). Content focused on teaching behavioural strategies and skills. |
| 14 | Flynn et al. (2018) | 442* | Male adolescents and adults with Borderline Personality Disorder and history of suicidal behaviour who receive treatment in community outpatient clinics | Protocol for a multi-site quasi- experimental study with non-equivalent groups | Ireland | Dialectical Behaviour Therapy (DBT) – adults received standard DBT over 12 months (individual and group sessions). Group skills delivered in 3 modules: mindfulness, distress tolerance, emotion regulation and interpersonal effectiveness. For adolescents, an adapted therapy (DBT-A) was delivered over 16 weeks. Modules adapted to be shorter and material made more developmentally appropriate. Additional module, *‘Walking the Middle Path’* and parents also included. |
| **No** | **Author (date)** | **n** | **Population/ sample** | **Study design** | **Country** | **Name and description of intervention** |
| 15 | Grey & McClean (2007) | 60 | Adults with intellectual disability and challenging behaviour | Non-randomised matched control group design | Ireland | Person Focused Training – staff trained for 9 days over a 6 month period and provided with skills in functional assessment and intervention development. |
| 16 | Griffith et al. (2016) | 20 | Adults with intellectual disability and aggression | Protocol for a single arm feasibility and qualitative study | UK | Using Mindfulness for Anger and Aggressive Behaviour with people with Learning Disabilities—Soles of the Feet (UMAA-LD SoF) – 6 sessions of mindfulness informed training taking up to 90 minutes delivered by a trained clinician. Included educational materials and home practice tasks. |
| 17 | Griffith et al. (2019) | 18 | People with intellectual disability (n=7), carers (n=6) and therapists (n=5) | Qualitative interviews | UK | See Griffith et al. (2016) above. |
| 18 | Hassiotis et al. (2018) | 246 | Adults with intellectual disability and challenging behaviour | Multicentre, single-blind, two-arm, parallel-cluster randomised controlled trial | UK | Positive Behavioural Support – manualised training to volunteer health staff in community services. Trained face-to-face over 6 days over the course of 15 weeks. Content included functional behavioural assessment, prevention and reactive strategies. It was advised that each participant should receive around 12.5 hours of intensive intervention. |
| 19 | Hoogsteder et al. (2016) | 26 | Adolescent/young adult (16-23) - offenders in an outpatient forensic setting | Quantitative experimental pre and post-test changes (pilot study) | The Netherlands | Responsive-Aggression Regulation Therapy (Re-ART) – cognitive behavioural based intervention that last between 5-18 months depending on level of risk. Sessions can be up to 3 times per week. |
| **No** | **Author (date)** | **n** | **Population/ sample** | **Study design** | **Country** | **Name and description of intervention** |
| 20 | Howells et al. (2000) | 5 | Adults with intellectual disability and aggression | Single case experimental and qualitative interviews | UK | Cognitive-Behavioural Anger Management training programme – 12 group sessions for 2 hours. This included training on aspects such as emotional recognition, physical and psychological signs of anger arousal, functional alternatives to aggression, etc. |
| 21 | Inchley-Mort & Hassiotis (2014) | 31 | Adults with intellectual disability (n=6), carers (n=25) | Semi-structured Qualitative interviews | UK | Complex Behaviour Service based on Positive Behavioural Support (PBS). Clinical psychologists, behavioural support worker and psychology graduate integrated within management structures and included in referral meetings over 12 months to provide an enhanced service and PBS interventions to manage challenging behaviour. |
| 22 | Inchley-Mort et al. (2014) | 46 | Adults with mild intellectual disability with complex behaviour  (control group n=22) | Observational study | UK | See Inchley-Mort & Hassiotis (2014) above. |
| 23 | Jones & Hollin (2004) | 8 | Adults with a diagnosis of personality disorder detained in a high security psychiatric hospital | Preliminary descriptive study | UK | "Managing Problematic Anger" treatment programme – 36 week manualised cognitive-behavioural programme targeting change in cognitive, arousal and behavioural aspects of anger. Included weekly individual and 2 hour group sessions. Delivered by nursing staff. |
| 24 | Jones et al. (2007) | 10 | Adolescents and adults with a dual diagnosis of developmental disability and psychiatric disorder | Experimental design (pilot study) | Canada | Community Anger Management Group – 12 2-hour sessions delivered by experienced clinicians and a doctoral student. Included education, skill-acquisition and behavioural rehearsal to address themes such as emotional identification, behavioural and cognitive coping strategies and arousal reduction. Included homework tasks. |
| **No** | **Author (date)** | **n** | **Population/ sample** | **Study design** | **Country** | **Name and description of intervention** |
| 25 | Karlin et al. (2014) | 71 | Veterans diagnosed with dementia and challenging behaviour in nursing home settings | Quantitative experimental multisite design | USA | STAR-VA - multicomponent psychosocial approach to managing challenging dementia-related behaviours. Manualised individually delivered intervention consisting of identifying and changing activators/consequences of behaviour, increasing meaningful events and promoting effective communication. Length of intervention depended on severity of behaviour (ranged from 4-147 days). |
| 26 | Khalid-Khan et al. (2016) | 7 | Adolescents with Borderline personality disorder or traits | Quantitative experimental. Within subjects pre-test to post-test | Canada | Dialectical Behaviour Therapy (DBT) modified for adolescents – 15-week course with 2.5 hour sessions led by two therapists. Adaptations included more time to check-in and reflect on use of coping skills, additional time for peer discussion and a psychodynamic component to examine early experience and trauma. |
| 27 | King et al. (1999) | 11 | Adults with mild intellectual disability and aggression | Quantitative experimental. Within subjects pre-test to post-test | Australia | Cognitive-Behavioural Anger Management intervention – 15 weekly group sessions (90 minutes). Explored the early signs of anger, coping skills training and behavioural rehearsal. |
| 28 | Klaver et al. (2020) | 24 | Staff members of 11 adults with intellectual disability and challenging behaviour | One group double pre-test to post-test design | The Netherlands | Positive Behavioural Support – 8 180-minute sessions every 2 weeks. Total training lasted 17 weeks. Training for staff included functional behaviour assessments, creating support plans with individualised goals and behavioural management techniques. |
| 29 | Kunik et al. (2020) | 228 | Individuals with Dementia with pain, depression or caregiver relationship problems and caregiver dyads | Randomised controlled trial | USA | Anger Prevention Training (APT) – skills-based intervention delivered over 6-8 weeks by licensed providers with behavioural health expertise and a telephone wrap-up session, each lasting 45 minutes. Core sessions included education, identification and management of pain, improving communication and increased pleasant activity planning. This was followed by 2-4 elective sessions to suit individual needs. Intervention primarily directed to caregiver. |
| **No** | **Author (date)** | **n** | **Population/ sample** | **Study design** | **Country** | **Name and description of intervention** |
| 30 | Lindsay et al. (2003) | 6 | Adults with intellectual disability convicted of violent personal assaults | Single case design with repeated measures | UK | Modified Anger Management Training – group sessions conducted weekly for 9 months (approx. 40 sessions) lasting 40-60 minutes. Incorporated cognitive restructuring and arousal reduction. |
| 31 | Lindsay et al. (2004) | 47 | Adults with mild intellectual disability | Experimental waitlist control design | UK | Anger Management Training – group sessions conducted weekly for 9 months (approx. 40 sessions) lasting 40-60 minutes. Incorporated arousal reduction, cognitive restructuring, problem solving and stress inoculation. |
| 32 | MacMahon et al. (2015) | 11 | Adults with intellectual disability | Qualitative interviews | UK | CBT Anger Management – group based intervention comprising of 12 sessions lasting for approximately 2 hours. Sessions focused on triggers, physiological and behavioural components of anger and behavioural and cognitive strategies. Delivered by staff within services. |
| 33 | Martin et al. (1998) | 27 | Adults with severe and profound intellectual disability and challenging behaviour | Double crossover quantitative experimental design | UK | Multi-Sensory Environment (MSE) – participants received blocks of MSE sessions and control sessions. The MSE sessions involved spending 1 hour in a multi-sensory room twice a week for 16 weeks. This had white walls, soft lighting, calming noises and sensory toys/objects. |
| 34 | McGill et al. (2018) | 81 | Adults with intellectual disability in social care settings | Pragmatic, cluster randomised controlled trial | UK | Positive Behavioural Support intervention – Researchers observed social care practice within services and drafted improvement plans to be implemented. This was supported through coaching and monthly monitoring over 8-11 months. |
| 35 | McWilliams et al. (2013) | 5 | Adults with mild to moderate intellectual disability, emotion regulation difficulties and challenging behaviour | Single case experimental design | New Zealand | Transformers Programme – community based group intervention to address emotion regulation difficulties and to teach coping skills. Delivered by at least two facilitators (service staff) in weekly sessions over a 6 month period. |
| **No** | **Author (date)** | **n** | **Population/ sample** | **Study design** | **Country** | **Name and description of intervention** |
| 36 | McWilliams et al. (2014) | n/a | People with intellectual disability convicted of an offence or have complex behaviour needs. | Descriptive paper detailing rationale and development | New Zealand | See McWilliams et al. (2013) above. |
| 37 | Neacsiu et al. (2014) | 101 | Female adults with a diagnosis of Borderline Personality Disorder and suicidal or self-injurious behaviour | Randomised controlled trial (secondary analysis) | USA | Dialectical Behaviour Therapy – cognitive-behavioural treatment delivered weekly for 1 year. Targeting life-threatening behaviours, behaviours that intervene with treatment delivery and quality of life. Included individual psychotherapy (1 hour a week) and groups skills training (2.5 hours a week) and phone consultations as needed. |
| 38 | Ong et al. (2019) | 72 | Children (6-12 years), with diagnoses of Disruptive Behavioural Disorders (n=35)  And typically developing children (n=37) | Quantitative experimental (pilot study) | Singapore | RegnaTales - series of 6 mobile app games designed to help children manage anger, based on a cognitive-behavioural framework. Children played the games for 50 minutes. Includes identifying feelings, coping skills and cognitive re-structuring. |
| 39 | Pearce et al. (2016) | 70 | Adults with personality disorders | Randomised controlled trial | UK | Democratic Therapeutic Community treatment – consisted of attendance at a preparatory group for 2 hours a week for up to a year. Participants were then able to join the group treatment via a democratic selection process and therapy was received for a maximum of 18 months. Therapy included shared decision making, reality confrontation and communalism. |
| 40 | Pert et al. (2013) | 15 | Adults with borderline to mild intellectual disability, psychiatric problems and aggression | Qualitative interviews | UK | Cognitive Behavioural Therapy – individually received sessions for emotional problems for 1 hour weekly or fortnightly. Participants received 10 sessions. |
| **No** | **Author (date)** | **n** | **Population/ sample** | **Study design** | **Country** | **Name and description of intervention** |
| 41 | Reynolds et al. (2019) | 442 | Children and adolescent psychiatric inpatients with various conditions (e.g. depression, anxiety, psychotic disorder) | Naturalistic prospective study using a pre-post design | USA | Modified version of Positive Behavioural Interventions and Supports (M-PBIS) – including reinforcement systems, problem-solving conversations and functional behaviour assessments. Implemented by trained hospital staff over a 26-month period and patients attended the programme for two weeks. |
| 42 | Rose (1996) | 5 | Adults with moderate to severe intellectual disability and aggression | Single case design | UK | Anger Management Group – 16 sessions lasting 1.5 hours. All participants were accompanied by a care worker. Sessions focused on identifying emotions, exploring appropriate and inappropriate responses to anger (using role-plays), improving self-awareness of cognitions and thought-stopping techniques. Included a self-monitoring diary. |
| 43 | Rose et al. (2005) | 86 | Adults with mild to moderate intellectual disability and aggression | Experimental design (with waiting list control) | UK | See Rose (1996) above. |
| 44 | Rose et al. (2008) | 41 | Adults with intellectual disability and aggression | Experimental  Pre-post-test design (with waiting list control) | UK | Cognitive-Behavioural Anger Management intervention – manualised individually delivered intervention focusing on emotional recognition, causes and manifestation of anger, problem solving and coping/prevention strategies. Flexibility in the provision of sessions (30-60 mins, between 14-18 sessions, usually weekly). Overall treatment length around 3-4 months. |
| 45 | Rose et al. (2009) | 62 | Adults with mild to moderate intellectual disability and aggression | Wait list design (n=23 in group intervention, n=18 in individual therapy and n=21 in waitlist control) | UK | Group and Individual Cognitive Behavioural interventions for Anger – 16 group sessions lasting 2 hours. Group content involved role plays of aggressive and other responses, thought-stopping and using positive self-statements. Content and delivery of individual sessions is explained in Rose et al. (2008) above. |
| **No** | **Author (date)** | **n** | **Population/ sample** | **Study design** | **Country** | **Name and description of intervention** |
| 46 | Rose et al. (2015) | 7 | Service managers in a day setting for people with intellectual disability | Qualitative study (as part of a randomised controlled trial) | UK | CBT Anger Management – manualised group based intervention comprised of 12 sessions, lasting 2 hours. Session content included triggers, physiological and behavioural components of anger, coping and cognitive/behavioural strategies. |
| 47 | Savage et al. (2004) | 8 | Male adults with dementia or schizophrenia on a geriatric psychiatric ward | Experimental design (within subjects pre- to post-test) | Canada | Agitation Management Model – included agitation assessments and discussions any time a participant exhibited agitated behaviour. Also included the planning and implementation of psychosocial interventions to prevent violence. Implemented by trained nursing staff. Assessments conducted at 12-week intervals for 36 weeks. |
| 48 | Singh et al. (2007) | 4 | 4 mother-child dyads, Mothers of children (aged 4-6) with developmental disabilities | Single case: Multiple baseline design | USA | Mindfulness Parent Training– individually delivered training for 12 weeks (2 hour sessions). Parents were taught meditation methods and were given exercises to practice mindfulness with their child. Mothers were asked to continue mindfulness practice for 52 weeks. |
| 49 | Singh et al. (2009) | 43 | Staff members (n=23) and individuals with intellectual disability (n=20) | Multiple baseline design | USA | Mindfulness Staff Training Programme – group delivered training for 12 weeks (2 hour sessions). Staff taught meditation methods and exercises to enhance mindfulness. Following training, the mindfulness practice phase lasted 40 weeks. |
| 50 | Singh et al. (2015) | 12 | Direct care professionals (n=9) and adults with developmental disabilities (n=3) | Single case: Multiple baseline design | USA | Mindfulness-Based Positive Behavior Support (MBPBS) – delivered as an intensive 5 day programme to staff. Training included meditation on the soles of the feet and how to use positive behaviour support within the context of mindfulness practices. The practice phase lasted between 32-37 weeks. |
| 51 | Singh et al. (2017) | 3 | Adolescents boys with Prader-Willi syndrome and their carers | Single case: Multiple baseline design | USA | Soles of the Feet Mindfulness – parents were trained over 4 weeks (total of 6 hours) and were instructed to use it during emotionally arousing situations. They then taught their child the procedure and encouraged them to practice this daily for 33-37 weeks. |
| **No** | **Author (date)** | **n** | **Population/ sample** | **Study design** | **Country** | **Name and description of intervention** |
| 52 | Singh et al. (2018) | 3 | Adults with a diagnosis of Alzheimer’s disease | Single case: Multiple baseline design | USA | Meditation on the Soles of the Feet – Trained during 30-min individual sessions (week 1) and 15-min sessions (weeks 2-4), five days a week. Participants taught a self-management mindfulness procedure. Instructed to then practice for 36-41 weeks to manage aggression. |
| 53 | Singh et al. (2020) | 123 | Adult carers from community group homes for people with mild-moderate intellectual disability | Randomised controlled trial | USA | Mindfulness-Based Positive Behavior Support (MBPBS) – training delivered in groups to caregivers over 10 weeks, followed by 30 weeks of intervention. Caregivers were taught different types of meditation and positive behavioural support practices. |
| 54 | Surr et al. (2020) | 675 | Adults with Dementia living in care homes | Pragmatic, cluster randomised controlled trial, open-cohort design | UK | Dementia Care Mapping – staff members were trained during a 4 day course to reduce agitation in care home residents through mapping observations, data analysis, reporting, monitoring and developing actions plans to improve the delivery of person-centred care. This was carried out in 3 cycles. |
| 55 | Taylor et al. (2002) | 20 | Adult male offenders with mild-borderline intellectual disability and aggression | Pilot study Delayed waitlist control design | UK | Cognitive Behavioural Therapy – manualised intervention consisting of a psychoeducation ‘preparatory phase’ to build rapport, and a ‘treatment phase’ including cognitive re-structuring, arousal reduction and behavioural skills training. 18 sessions of individual treatment were delivered twice weekly over 12 weeks. |
| 56 | Taylor et al. (2005) | 40 | Adult offenders with mild-borderline intellectual disability and aggression | Delayed waitlist control design | UK | See Taylor et al. (2002) above. |
| **No** | **Author (date)** | **n** | **Population/ sample** | **Study design** | **Country** | **Name and description of intervention** |
| 57 | Tournier et al. (2020) | n/a | Individuals with intellectual disability and challenging behaviour | Developing a logic model | The Netherlands | Triple C (Client, Coach, Competence) intervention- values driven framework that includes involvement at all levels within an organisation. The core focus is for person and support workers to build an unconditional supportive relationship, increase competencies and carry out meaningful activities together. |
| 58 | Wetterborg et al. (2020) | 30 | Adult males with Borderline Personality Disorder, antisocial behaviour and criminal behaviours in community outpatient settings | Within-group design with repeated measures | Sweden | Dialectical Behaviour Therapy (DBT) – participants received DBT over 12 months from a clinical psychologist or psychiatric nurse. This consisted of 1 hour of individual therapy and 2.5 hours of group skills training per week. Minor adaptations were made to the standard DBT format. An additional teaching validation module was added and skills’ training was delivered at a slower pace. |
| 59 | Willner et al. (2013) | 179 | Adults with intellectual disability | Cluster randomised controlled trial | UK | Cognitive Behavioural Therapy Anger Management Intervention – manualised intervention comprising of 12 weekly 2 hour group sessions for day service staff. Intervention content prioritised behaviour change and coping skills. |
| *anticipated n | | | | | | |
