## Supplementary Materials 3 for "Complex interventions for aggressive challenging behaviour in adults with intellectual disability: a rapid realist review informed by multiple populations"

**Supplementary Table 3.** Relevance and rigour judgements as a means of quality appraisal

| Study ID | Relevance | | | Rigour | | |
| --- | --- | --- | --- | --- | --- | --- |
|  | Judgement | | Reasoning | | Judgement | Reasoning |
| Singh et al. (2017) | ✓ | | A | | ✓ | MMAT score: 60 |
| Cullen et al. (2012) | ✓ | | A | | 🗶 | ROB2 score: High risk |
| Kunik et al. (2020) | 🗶 | | -A, -B | | 🗶 | ROB2 score: High risk |
| McWilliams et al. (2014) | ✓ | | A, B | | 🗶 | MMAT score: 20 |
| Ong et al. (2019) | 🗶 | | -A, -B | | ✓ | MMAT score: 60 |
| Lindsay et al. (2004) | ✓ | | A, B | | 🗶 | ROB2 score: High risk |
| Jones et al. (2007) | 🗶 | | -A, -B | | ✓ | MMAT score: 60 |
| Rose (1996) | ✓ | | A, B | | 🗶 | MMAT score: 40 |
| Martin et al. (1998) | ✓ | | B | | ✓ | MMAT score: 80 |
| Ballard et al. (2009) | ✓ | | A | | ✓ | MMAT score: 80 |
| Griffiths et al. (2016) | ✓ | | B | | 🗶 | Appraisal tools inappropriate |
| Pert et al. (2013) | ✓ | | B | | ✓ | CASP score: 17 |
| King et al. (1999) | ✓ | | B | | ✓ | MMAT score: 80 |
| Taylor et al. (2002) | 🗶 | | -A, -B | | ✓ | ROB2 score: some concerns |
| Singh et al. (2020) | ✓ | | A, B | | ✓ | ROB2 score: some concerns |
| Inchley-Mort et al. (2014) | ✓ | | B | | ✓ | MMAT score: 100 |
| Savage et al. (2004) | 🗶 | | -A, -B | | ✓ | MMAT score: 60 |
| Pearce et al. (2017) | 🗶 | | -A, -B | | ✓ | ROB2 score: low risk |
| Tournier et al. (2020) | 🗶 | | -A, -B | | 🗶 | Appraisal tools inappropriate |
| Wetterborg et al. (2020) | ✓ | | A | | 🗶 | ROB2 score: High risk |
| Flynn et al. (2018) | ✓ | | A | | 🗶 | Appraisal tools inappropriate |
| Khalid-Khan et al. (2016) | 🗶 | | -A, -B | | 🗶 | MMAT score: 40 |
| Appelhof et al. (2019) | ✓ | | A | | ✓ | MMAT score: 60 |
| Borowsky et al. (2004) | ✓ | | A | | ✓ | ROB2 score: low risk |
| Benson et al. (1986) | ✓ | | B | | ✓ | MMAT score: 80 |
| Ahemaitijiang et al. (2020) | ✓ | | A | | 🗶 | MMAT score: 40 |
| Singh et al. (2015) | ✓ | | A | | ✓ | MMAT score: 60 |
| Howells et al. (2000) | ✓ | | B | | 🗶 | MMAT score: 20 |
| Rose et al. (2005) | ✓ | | B | | ✓ | ROB2 score: some concerns |
| Klaver et al. (2020) | ✓ | | A, B | | ✓ | MMAT score: 80 |
| Rose et al. (2009) | ✓ | | B | | ✓ | ROB2 score: some concerns |
| Neacsiu et al. (2014) | ✓ | | A | | ✓ | ROB2 score: low risk |
| Bradshaw et al. (2004) | ✓ | | B | | ✓ | MMAT score: 60 |
| Reynolds et al. (2019) | ✓ | | A | | ✓ | MMAT score: 100 |
| Rose et al. (2008) | ✓ | | A, B | | ✓ | ROB2 score: some concerns |
| Lindsay et al. (2003) | ✓ | | A, B | | ✓ | MMAT score: 60 |
| Jones & Hollin (2004) | ✓ | | A | | ✓ | MMAT score: 60 |
| Singh et al. (2019) | ✓ | | A | | ✓ | MMAT score: 60 |
| Singh et al. (2007) | 🗶 | | -A, -B | | 🗶 | MMAT score: 20 |
| Singh et al. (2009) | ✓ | | B | | ✓ | MMAT score: 80 |
| Griffith et al. (2019) | ✓ | | B | | ✓ | CASP score: 16 |
| Bambara et al. (2001) | ✓ | | A | | ✓ | CASP score: 16 |
| Karlin et al. (2014) | 🗶 | | -A, -B | | ✓ | MMAT score: 80 |
| Edwards et al. (2019) | ✓ | | A | | ✓ | MMAT score: 80 |
| Grey & McClean (2007) | ✓ | | A | | ✓ | MMAT score: 80 |
| Chilvers et al. (2011) | ✓ | | A | | ✓ | MMAT score: 60 |
| Hoogsteder et al. (2016) | ✓ | | A | | 🗶 | MMAT score: 40 |
| McWilliams et al. (2014) | ✓ | | A, B | | 🗶 | Appraisal tools inappropriate |
| Taylor et al. (2005) | ✓ | | A | | 🗶 | ROB2 score: High risk |
| Study ID | Relevance | | | | Rigour | |
|  | Judgement | | Reasoning | | Judgement | Reasoning |
| Inchley-Mort & Hassiotis (2014) | ✓ | | A, B | | ✓ | CASP score: 17 |
| Hassiotis et al. (2018) | ✓ | | B | | ✓ | ROB2 score: low risk |
| McGill et al. (2018) | ✓ | | A, B | | ✓ | ROB2 score: low risk |
| Bowers et al. (2015) | ✓ | | A | | ✓ | ROB2 score: low risk |
| Davies et al. (2020) | ✓ | | A | | ✓ | MMAT score: 60 |
| Rose et al. (2015) | ✓ | | A, B | | ✓ | CASP score: 17 |
| MacMahon et al. (2015) | ✓ | | A, B | | ✓ | CASP score: 19 |
| Willner et al. (2013) | ✓ | | A, B | | ✓ | ROB2 score: low risk |
| Surr et al. (2020) | 🗶 | | -A, -B | | ✓ | ROB2 score: low risk |
| Ballard et al. (2020) | 🗶 | | -A, -B | | ✓ | ROB2 score: some concerns |
| ✓ | *Record deemed more relevant/more rigorous* | | | | | |
| 🗶 | *Record deemed less relevant/less rigorous* | | | | | |
| A | *Record contributed significantly to theory building* | | | | | |
| B | *Record recruited a sample of adults with ID receiving an intervention in the community* | | | | | |
| -A, -B | *Record did not meet either condition A or condition B above* | | | | | |
